# Coverage and correlates of COVID-19 vaccination among children aged 5-11 years in Alberta, Canada

**DOI:** 10.1101/2022.08.29.22279354

**Authors:** Shannon E. MacDonald, Laura Reifferscheid, Yuba Raj Paudel, Joan Robinson

## Abstract

**Background and Objectives:** In Alberta, Canada, the COVID-19 vaccination program for children aged 5-11 years opened on November 26, 2021. Our objectives were to determine the cumulative vaccine coverage, stratified by age, during the first seven months of vaccine availability, and investigate factors associated with vaccine uptake.

**Methods:** This retrospective cohort study used population-based administrative health data to assess COVID-19 vaccination coverage among children aged 5-11 years in Alberta, Canada. We determined cumulative vaccine coverage since the time of vaccine availability and used a modified Poisson regression to evaluate factors associated with vaccine uptake.

**Results:** Of 377,753 eligible children, 43.8 % (n=165,429) received one or more doses of COVID-19 vaccine during the study period (11.2% received only one dose, while 32.5 % received 2 doses). Almost 90% of initial doses were received within the first two months of vaccine availability. Of those eligible for a second dose, only 75.1% (n=122,973) received it during the study time period. We found a step-wise relationship between increasing child age and higher vaccine coverage. Other factors associated with higher vaccine coverage included living in a neighborhood with higher income, in a more densely populated area, and in certain geographic health zones. Registration in a private school was associated with lower vaccine coverage.

**Conclusions:** Messaging around COVID-19 vaccine safety and need should be tailored to child age, rather than uniform across the 5-11 year age range. Opportunities for targeted vaccination interventions should be considered.

## Introduction

Health Canada approved the first COVID-19 vaccine product for use among 5-11 year old children in Canada on November 19, 2021.^1^ As of July 17, 2022, approximately 56% of Canadian children aged 5-11 had received at least one dose.^2^ While coverage increased rapidly during the first two months of vaccine availability, uptake has largely stalled since that time,^2^ potentially indicating that current vaccination strategies have reached saturation in the population.

The success of childhood vaccination programs is dependent on the willingness of parents to obtain the vaccine for their children, supported by acceptable and equitable vaccination delivery. Previous studies have explored parental intentions toward COVID-19 vaccination for their children, in anticipation of vaccine offerings. A global pooled estimate compiled in December 2021 reported that approximately 60% of parents intended to vaccinate their children, while approximately 22.9% intended to refuse vaccination.^3^ However, results varied widely, and studies that provided for uncertainty in decision making indicated approximately 26% remained undecided about vaccinating their children.^3^ Logistical or structural barriers caused by inaccessible or inconvenient vaccination delivery can exacerbate these uncertainties, and prevent parents from acting on positive vaccination attitudes^4^

These factors, along with rapidly changing pandemic conditions, and continuously increasing amounts of information and misinformation about the COVID-19 vaccine, means that context-specific COVID-19 vaccine uptake among children should be closely monitored. Although COVID-19 vaccine coverage surveillance estimates are available from various government sources, there is little published research exploring uptake patterns among children aged 5-11 years, including variability in coverage across the age spectrum. Measuring age-specific COVID-19 vaccine coverage, and determining associated factors, are necessary to ensure vaccination programs are effectively tailored to population needs. Thus, we sought to: 1) estimate coverage of COVID-19 vaccine among children aged 5-11 years in Alberta, 2) assess the increase in vaccine coverage over time since vaccine availability, and 3) investigate factors associated with vaccination.

## Methods

We conducted a retrospective cohort study of COVID-19 vaccine coverage and correlated factors using linked administrative health data in Alberta, Canada. Vaccination status was assessed between the start of the COVID-19 vaccine program for 5-11 year olds (November 26, 2021) and the end of the study period (June 26, 2022).

### Setting

Alberta has a population of approximately 4.4 million, and is divided into five geographic health zones. During our study period, the only COVID-19 vaccine product available to children 5-11 years was BNT162b2 (COMIRNATY by Pfizer). Doses were offered mainly through public health centres, as well as a small number of pharmacies in areas where public health centres were not nearby.^5^ Although some vaccination requirements were put in place for children 12-17 years in some jurisdictions (some youth extracurricular activities required vaccination),^6^ no such restrictions were placed on younger children.^7^

### Cohort and Data Sources

More than 99% of Albertans are registered with the province’s universal healthcare insurance program. Once registered, each individual is provided a personal healthcare number, which acts as a unique lifetime identifier. Using this identifier, we deterministically linked administrative databases available through the Alberta Ministry of Health. Through the school registration database (Provincial Student Immunization Repository [PSIR]), we identified all children enrolled in Alberta schools during the 2021/2022 school year, as well as individual birth date, sex, and postal code of residence. Children were included in the cohort if they became eligible for vaccination on November 26, 2021 (i.e. date of birth between January 01, 2010 and November 26, 2016, aged 5-11 years; children born in 2009 were already eligible for vaccination as a 12 year old ^8^). We used a Postal Code Conversion File to link postal codes to Statistics Canada’s 2016 census information, to determine place of residence (by population density) and neighbourhood income quintile. Individual vaccination status was retrieved from the provincial Immunization and Adverse Reaction to Immunization (Imm/ARI) repository, which includes all COVID-19 vaccine doses administered, regardless of provider. Children who died or departed from Alberta between September 01, 2021 (the start of the school year) and June 26, 2022 (end of study period) were excluded from the analysis. We excluded children with a residential postal code from outside Alberta, or in the city of Lloydminster (where vaccinations are provided by the neighboring province). Children with a two-dose interval less than the minimum recommendation of 21 days ^8^ were excluded, as either dose may have represented a data entry error. Finally, those who received a COVID-19 vaccine dose before the vaccine was available in Alberta (November 26, 2021) were excluded, as doses may represent a data entry error, and to ensure uniform follow-up time for all children to calculate cumulative coverage. See Figure A1 for a flowchart of cohort selection.

### Measures and Analysis

To identify vaccine uptake trends over time, we calculated cumulative one- and two-dose vaccine coverage overall, and by age, for the study period. To calculate cumulative coverage, we divided the daily number of children who received a first or second vaccine dose by the total population of children. We calculated the absolute difference in cumulative coverage between two time points to measure increase in coverage. Vaccine delivery location was categorized into public health centres, pharmacies, physicians, and others.

We categorized vaccination status into four types, based on number of doses and vaccination timing: (1) No doses - no COVID-19 vaccine doses received during our study period; (2) One dose dropouts - eligible for two doses within our study period but only received one dose (received first dose between November 26, 2021 and May 01, 2022; therefore, eligible for a second dose during study period based on recommended two-dose interval of 8 weeks^8^); 3) One dose late acceptors - received a first dose but were not yet eligible for a second dose during our study period (received first dose after May 1, 2022); and, 4) Two doses - received two doses during our study period.

To compare the characteristics of children who received at least one dose of vaccine to those who received none, we calculated frequencies and percentages for six exposure variables of interest: age as of November 26, 2021; sex; place of residence by population density (metro, moderate metro, urban, moderate urban, rural centre, rural, and rural remote^9^); geographic health zone (South, Calgary, Central, Edmonton, and North); neighborhood income quintile; school authority type (Public, Publicly-funded Catholic, Private, Charter, Francophone, and early childhood services [ECS] private operator [education programs for children under the age of 6]). Children registered for homeschooling are registered with the relevant school authority in Alberta; thus, are included in these groups.^10^ A sub-group analysis was conducted among children aged 11 years at the start of the study period to assess if vaccine coverage differed between those who turned 12 years during the study period and those who remained 11.

Using a modified Poisson regression analysis, we determined the adjusted risk ratio (ARR) and 95% confidence interval (CI) for the relationship between vaccination status and each exposure variable of interest (age, sex, place of residence, health zone, neighborhood income quintile, and school authority type). Data analysis was completed using SAS version 9.4. Ethics approval for this work was obtained from the University of Alberta’s Health Research Ethics Board.

## Results

A total of 377,753 children were included in our cohort. Demographic characteristics of the cohort are provided in Table 1.

**Table 1.** Cohort characteristics (N=378,355)

| Characteristic | n | % |
| --- | --- | --- |
| **Age<sup>a</sup>** |  |  |
| 5 years | 50,980 | 13.5 |
| 6 years | 55,112 | 14.6 |
| 7 years | 55,806 | 14.8 |
| 8 years | 54,756 | 14.5 |
| 9 years | 55,495 | 14.7 |
| 10 years | 55,268 | 14.6 |
| 11 years | 50,336 | 13.3 |
| **Sex** |  |  |
| Female | 183,983 | 48.7 |
| Male | 193,599 | 51.3 |
| Missing | 171 | 0.1 |
| **Income Quintile** |  |  |
| Q1 (lowest income) | 60,652 | 16.1 |
| Q2 | 66,132 | 17.5 |
| Q3 | 68,428 | 18.1 |
| Q4 | 86,758 | 23.0 |
| Q5 (highest income) | 95,783 | 25.4 |
| **Place of residence** |  |  |
| Metro | 198,119 | 52.5 |
| Moderate metro influence | 59,968 | 15.9 |
| Urban | 35,648 | 9.4 |
| Moderate urban influence | 11,380 | 3.0 |
| Rural centre area | 12,887 | 3.4 |
| Rural | 51,640 | 13.7 |
| Rural remote | 8,111 | 2.2 |
| **Geographic Health Zone** |  |  |
| Calgary | 144,371 | 38.2 |
| Edmonton | 124,197 | 32.9 |
| South | 27,619 | 7.3 |
| Central | 38,346 | 10.2 |
| North | 43,220 | 11.4 |
| **School authority type** |  |  |
| Public | 252,788 | 66.9 |
| Publicly-funded Catholic | 88,404 | 23.4 |
| Private School | 22,019 | 5.8 |
| Charter | 5,753 | 1.5 |
| Francophone | 5,569 | 1.5 |
| ECS Private Operator | 1,577 | 0.4 |
| Missing | 1,643 | 0.4 |

<sup>a</sup> Age at start of study period (Nov 26, 2021)

We observed a similar pattern of increasing coverage with time across the age groups, with a stepwise increase in vaccine coverage with increasing age (Figures 2 and 3). Within two months of vaccine availability, first dose coverage had reached more than 89% of end-of-study coverage for all age groups except the oldest (11 years of age at the start of the study period) (Figure 2, Table A1). Coverage in the oldest age group increased at a higher rate throughout the remainder of the study period, though the increase was less than 4% between January 26 and February 26, and less than 1% in subsequent months for all age groups. Receipt of one or more doses of vaccine was significantly higher among those who turned 12 during our study period, compared to those who remained 11 (55.5% *vs* 50.1%, p<0.0001) (Table A2).

**Figure 2.**
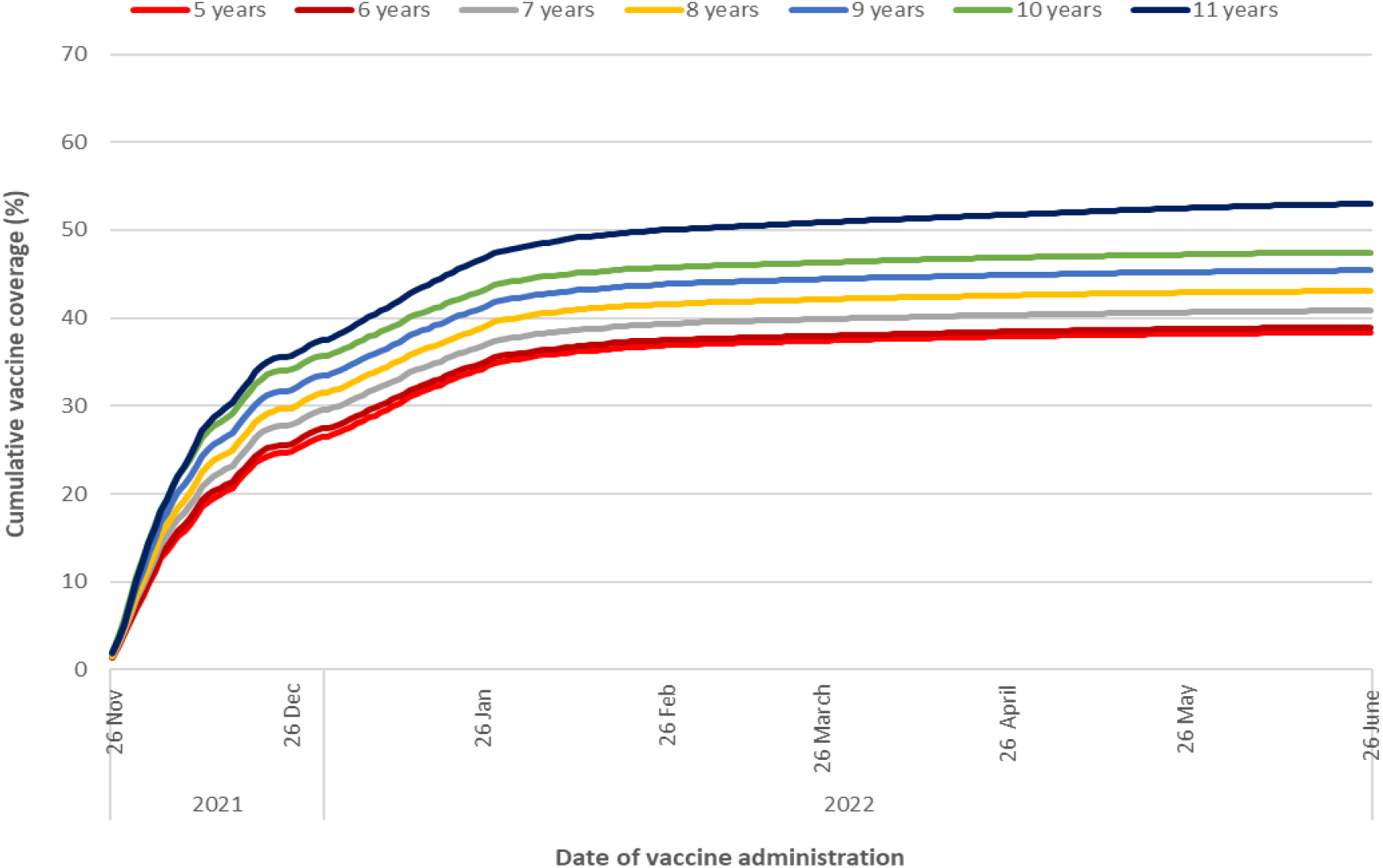
Cumulative coverage for one dose of COVID-19 vaccine by age category.

**Figure 3.**
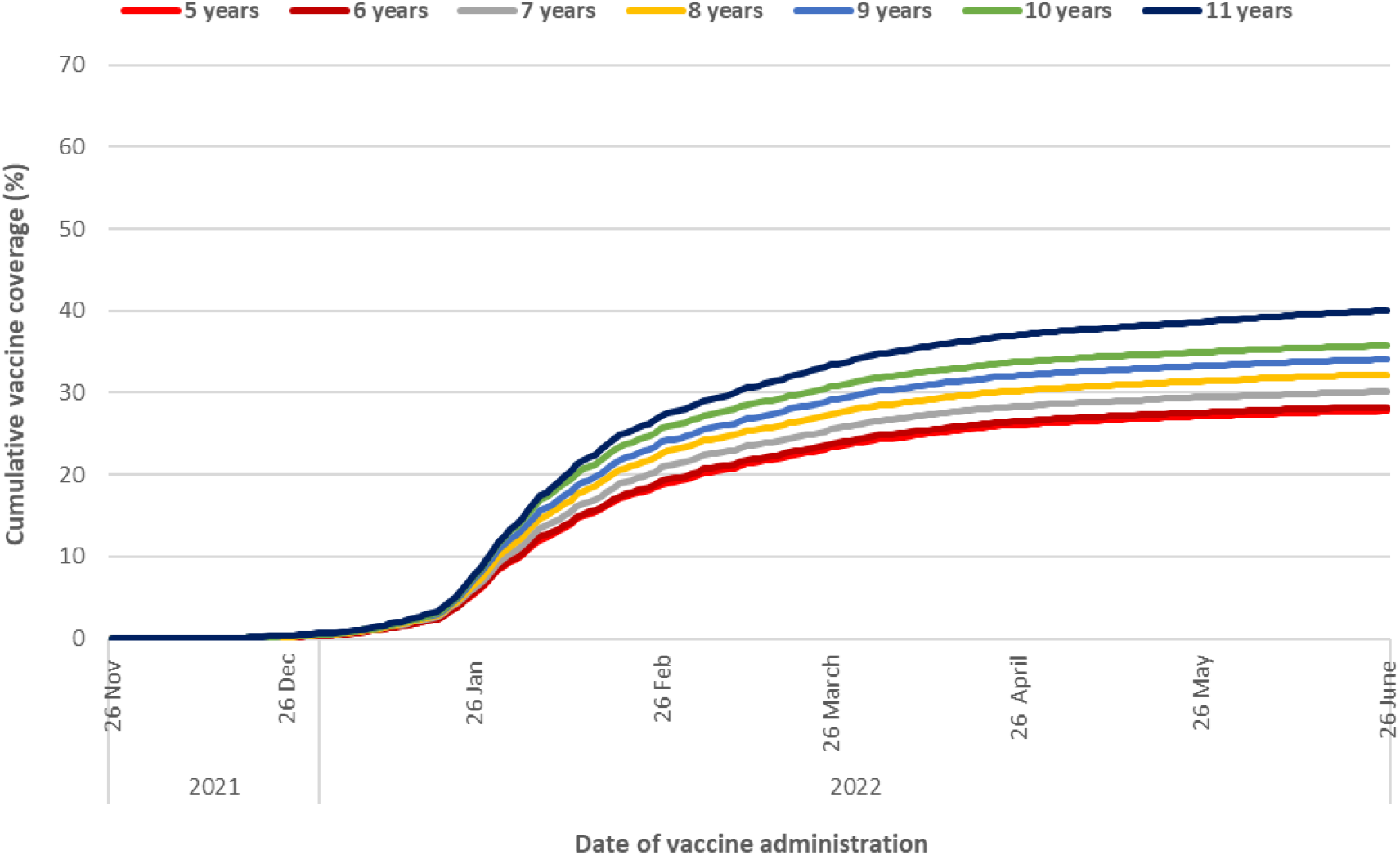
Cumulative coverage for two doses of COVID-19 vaccine by age. Recommended interval between first and second dose was 8 weeks Over half of the cohort received no vaccine doses (56.2%, n=212,324), while the remaining 43.8% (n=165,429) received one or more doses (Figure 4). Of the children who received one or more doses, most completed the two dose series during our study period. A total of 10.8% (n=40,716) of the cohort received only one dose of vaccine, despite being eligible for a second dose during the study period (Dropouts), representing 95.8% of the one dose recipients. No dose, one dose, and two dose coverage by study characteristics are presented in Table A4. Among those who received two doses, the average dose interval was 67 days (SD:22-24 days) for all age categories (Table A3).

A total of 43.8 % (n=165,429) of the cohort had received at least one dose by the end of our study period (11.2% received only one dose, while 32.5 % received 2 doses) (Figure 1). First dose vaccine coverage reached 30% within the first month of vaccine availability (by December 26, 2021), and increased another 9.1% in the second month (by January 26, 2022) (Figure 1, Appendix Table A1). During the third month of availability, first dose coverage increased by 3%, reaching 42.1% on February 26, 2022. For the remaining four months of follow-up, first dose vaccine coverage increased by less than one percent per month. Two-dose coverage reached 32.5% (n=122,973) by the end of the follow-up period. The majority of vaccine doses were distributed by Public Health centers (94.4%), with only 4.2% distributed by Pharmacies, and remaining doses (1.4%) distributed from other sources (including First Nations clinics and physician clinics).

**Figure 1.**
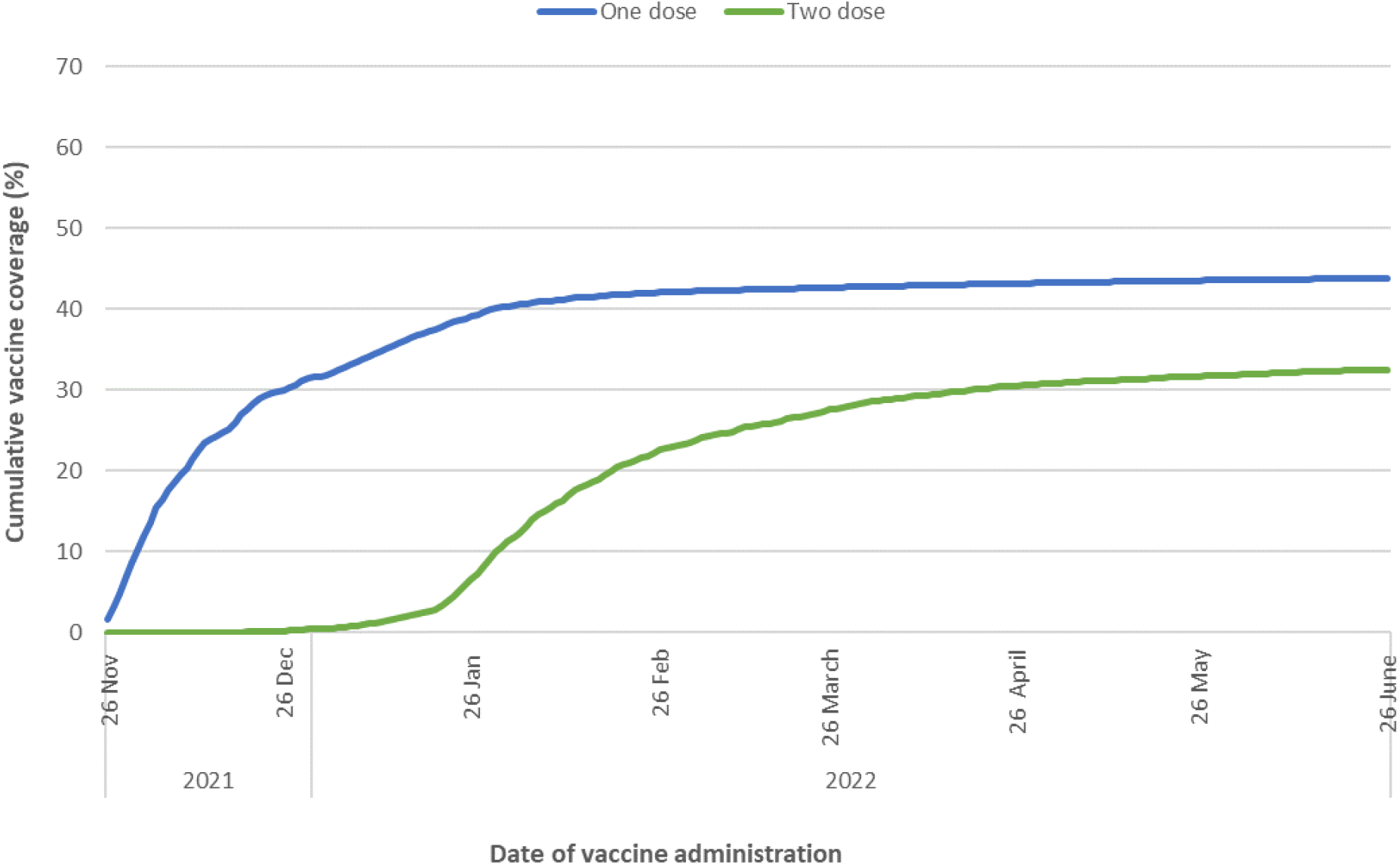
Cumulative percent of children who have received one and two doses of COVID-19 vaccine.

## Discussion

We used school enrollment records and a population-based immunization repository to determine COVID-19 vaccine coverage among children aged 5-11 years during the first seven months of vaccine availability in Alberta, Canada. Our results indicate less than half had received a dose of COVID-19 vaccine by June 26, 2022, and most had received their initial dose within the first two months of vaccine availability. Similar patterns in vaccine uptake have been noted in other Canadian provinces,^2^ and other countries^11^, despite different levels of coverage. Our coverage level was lower than previously reported parental intentions for similar age groups in Canada (56% intended, 33% undecided for children 5-11 years^12^) and Alberta (60% intended, 31% undecided for children aged 9-12 years^13^), suggesting that a significant portion of parents with unvaccinated children may still be considering vaccination.

Notably, coverage in Alberta is lower than other Canadian provinces and territories, which are estimated to range from 52.8 - 87.4% (as of June 19, 2022^2^). This discrepancy is not apparent for other childhood vaccines^14^, indicating that regional efforts to promote COVID-19 vaccine uptake should be explored. For example, school-based delivery of COVID-19 vaccines has been used in Quebec, Saskatchewan, and British Columbia.^12^ School-based programs have been used effectively in Canada to achieve high and equitable coverage for other childhood vaccines;^15,16^ in fact, simply encouraging vaccination in school may promote uptake among this age group.^17^ Our results also indicate that prioritizing vaccination interventions by school type may be useful for targeting areas of low coverage. However, lower vaccine coverage among those attending private schools has been reported for other routine vaccines.^18,19^ In addition, while Canadian parents who support COVID-19 vaccination for their children are largely in favor of school-based delivery, those who are undecided may be less supportive.^12^ The province of Alberta did expand delivery options in March 2022, increasing availability through pharmacies and primary care physician offices, and offering walk-in appointments and extended hours at select public health centres.^20,21^ These efforts did not appear to impact provincial-level vaccine coverage; however, as interventions were applied only in specific areas, there may have been significant localized impacts. In addition, these efforts may have been more effective if implemented earlier in the vaccine roll-out, when interest was high. More information about the challenges and successes of regional programs, school-based and otherwise, is required.

Nearly one quarter of children who received an initial dose of COVID-19 vaccine did not receive a second dose within the study timeframe, despite being eligible. These “dropouts” reflect a missed opportunity to increase vaccine series completion, which is crucial to ensure protection is maximized. Reminders for upcoming vaccination doses, and recalls for overdue vaccinations, have been identified as effective strategies for increasing vaccination rates among children;^22^ tailoring these interventions to the local population may be particularly effective.^23^ Additionally, techniques to alleviate first dose pain and anxiety may promote vaccine compliance, particularly in this age group.^24^ Given the variable rate in follow-up time after first dose receipt, we did not explore factors associated with delaying or not receiving a second dose. Future research should recognize this group as a unique population, and determine factors associated with incomplete and/or delayed vaccination.

In our study, younger age was significantly associated with lower vaccine coverage. This pattern persisted throughout the age range, with higher coverage even noted among children who turned 12 years during the study time period compared to those who remained 11. However, estimated coverage among those who turned 12 was still below estimated coverage for the 12-17 year old age group in Alberta (88%^2^), indicating that the relationship between age and uptake may extend to older children. Previous survey results indicate mixed results on the impact of child age on parental vaccination intention ^3^; however, studied age ranges varied. Our findings are consistent with a Canadian survey that reported parents of 8-11 year olds were more likely to report positive vaccination intentions than those of 5-7 year olds (58.1% *vs* 54.8%).^12^ Murthy et al (2022) found a similar relationship between age and coverage during the first two months of vaccine availability in the US; our results indicate that this pattern persists. Therefore, treating 5-11 year olds as one homogeneous group for messaging and vaccine delivery may not be appropriate for optimizing vaccine coverage. Instead, age-specific information on vaccine safety and effectiveness should be provided, including greater efforts to publicize the growing number of children worldwide who have been safely vaccinated. This may be particularly important for the large portion of parents who have stated they wish to “wait and see”.^3,25,26^

We found that vaccine coverage increased with both income and population density, and varied by geographic health zone. These relationships are consistent with patterns noted for adult COVID-19 vaccine intention and uptake ^27,28^, which is not surprising given that parental vaccination status is highly predictive of vaccination intention for their children.^29^ However, the vaccination rate for children in this age group is much lower than that reported for parental-age adults (approximately 90% ^2^). This disparity has also been noted for childhood vaccination intentions among parents,^12,29^ with lower intentions associated with perceptions of lower risk of severe COVID-19 disease in children^3^ and greater concern over unknown or potential side effects related to vaccination.^26^ This means that trusted healthcare providers must have the information and training required to have effective conversations in their communities.^30^ Efforts to promote COVID-19 vaccination for children 5-11 as a social norm by leveraging informal social connections may also be effective.^25,31^

### Strengths and limitations

This study benefited from timely population-based data, using individual-level school enrollment and immunization records. By using school enrollment records to identify our cohort we were able to accurately identify children who are current residents of Alberta. In addition, our overall coverage estimates were similar to those posted by the Alberta Ministry of Health, who apply population weights to measure coverage. However, we were only able to include the first seven months of vaccination data, limiting our ability to comment on longer-term trends. Ongoing analysis of COVID-19 vaccine coverage in this age group is required.

## Conclusions

Our results indicate that parents either sought vaccination for their 5-11 year old children soon after it was available or not at all. Thus, innovative messaging and delivery options will be required to increase uptake in this age group over the longer term. Although this uptake pattern is consistently noted across Canadian provinces, lower coverage levels in Alberta indicate that understanding regional differences in COVID-19 vaccination efforts is required. We found that child age was significantly related to vaccine coverage, even after correcting for sociodemographic and regional variability, suggesting that the 5-11 year old age group should not be treated as one homogeneous group. Rather, tailored messaging to parents of younger children within this age group should be used. Although our analysis was conducted only seven months after vaccine availability, we found some indication that efforts to promote timeliness of two-dose series completion are required. These important lessons should be used to guide COVID-19 vaccine programming for children 6 months to 4 years.

## Data Availability

The steward of the data used in this study is the Alberta Ministry of Health, who maintains the data for the purpose of health system administration. Thus, the authors are not at liberty to make the data publicly available.

## Ethical approval

The University of Alberta Research Ethics Board granted ethical approval for this study.

## Data Availability

The steward of the study used in this study is the Alberta Ministry of Health, who maintains the data for the purpose of health system administration. Thus, the authors are not at liberty to make the data publicly available.

**Figure 4.**
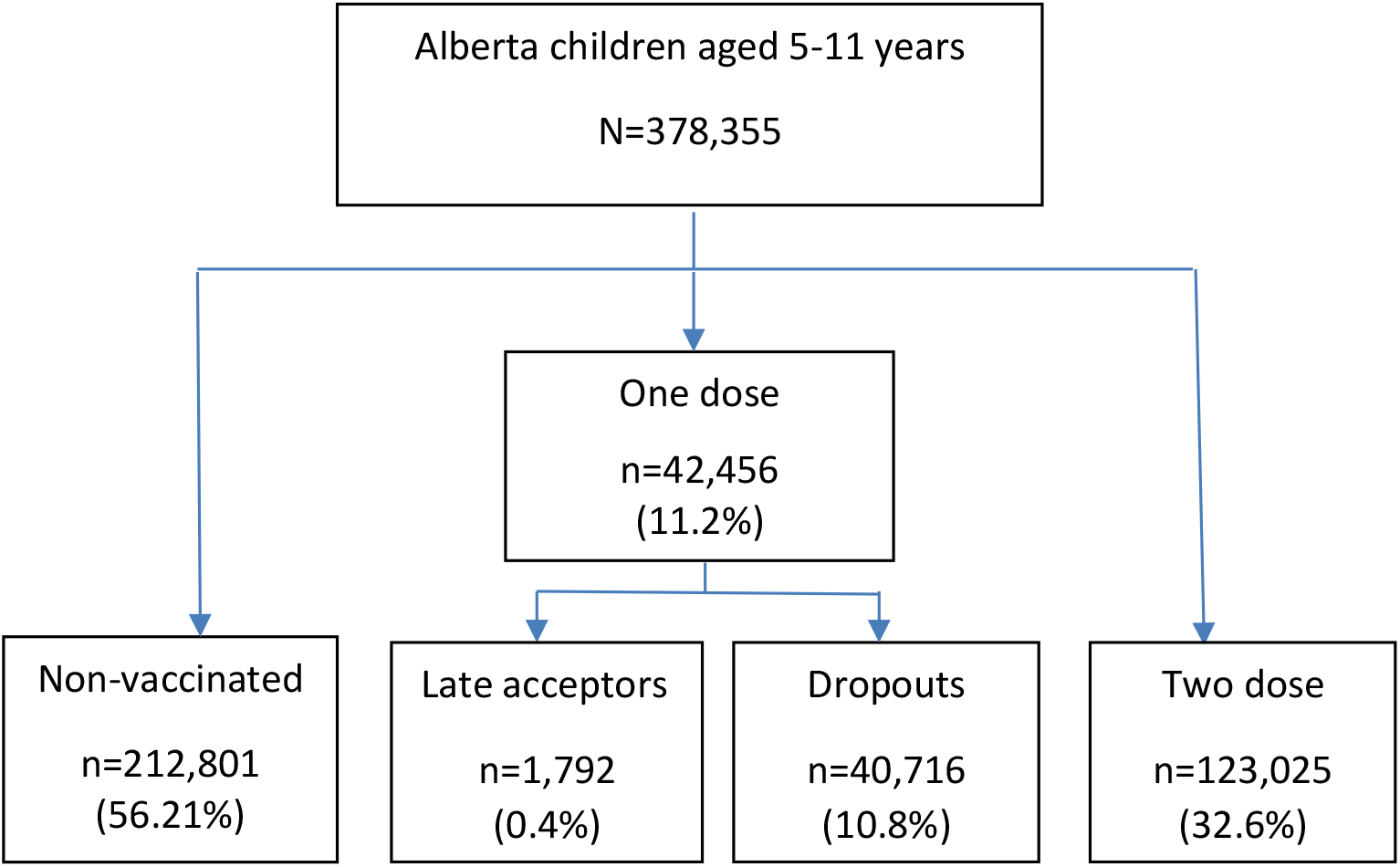
COVID-19 vaccine coverage by vaccination status category at end of follow-up period. No dose: received no doses; One dose, late acceptors: received one dose, were not time-eligible for a second dose; One dose, dropouts: received one dose, did not receive a second dose despite time-eligibility; Two dose: received two doses.

In the multivariable analysis, factors positively associated with receiving at least one dose of COVID-19 vaccination were older age (e.g. 11 years *vs* 5 years ARR 1.40 [1.38–1.42]; 10 years *vs* 5 years ARR 1.26 [1.24–1.27]); living in a higher income neighborhoods (e.g. highest quintile *vs* lowest quintile [ARR 1.82 [1.80–1.85]) (Table 3). By contrast, living in a moderate urban region (ARR 0.62 [0.59–0.65]), rural (ARR 0.63 [0.61–0.64]) or rural remote areas (ARR 0.68 [0.65–0.72]) were associated with lower vaccination compared to living in a metro areas. Similarly, living in the South (ARR 0.88 [0.85–0.91], Central (ARR 0.71[0.69-0.73]), and North health zones (ARR 0.59 [0.58–0.61] were associated with lower vaccination, compared to the Calgary zone. In comparison to those attending Public schools, children attending a Charter (ARR 1.18 [1.16–1.21]), or publicly-funded Catholic school (ARR 1.03 [1.02–1.04]) were more likely to be vaccinated, while those attending Francophone (ARR 0.95[0.93-0.98], Private (ARR 0.66 [0.64-0.67], and ECS private Operators (ARR 0.87 (0.82–0.93] were less likely to be vaccinated. No appreciable differences in coverage were observed by sex.

**Table 2.** Proportion of children who received at least one dose of COVID-19 vaccine, with unadjusted and adjusted risk ratios for factors associated with vaccination.

| Characteristic | Coverage ($\geq 1$ dose), % (n) | Unadjusted risk ratio (95% CI) | Adjusted risk ratio (95% CI) | p-value |
| --- | --- | --- | --- | --- |
| **Age (years)** |  |  |  |  |
| 5 years | 38.4 (19,553) | Ref | Ref |  |
| 6 years | 38.9 (21,446) | 1.01 (1.00, 1.03) | 1.03 (1.01, 1.04) | 0.0002 |
| 7 years | 40.8 (22,778) | 1.06 (1.05, 1.08) | 1.08 (1.06, 1.09) | <.0001 |
| 8 years | 43.1 (23,570) | 1.12 (1.11, 1.14) | 1.14 (1.13, 1.16) | <.0001 |
| 9 years | 45.4 (25,196) | 1.18 (1.17, 1.20) | 1.20 (1.19, 1.22) | <.0001 |
| 10 years | 47.5 (26,225) | 1.24 (1.22, 1.25) | 1.26 (1.24, 1.27) | <.0001 |
| 11 years | 53.0 (26,661) | 1.38 (1.36, 1.40) | 1.40 (1.38, 1.42) | <.0001 |
| **Sex** |  |  |  |  |
| Male | 43.6 (84,374) | Ref | Ref |  |
| Female | 44.0 (80,953) | 1.01 (1.00, 1.02) | 1.01 (1.00, 1.02) | 0.0155 |
| **Neighborhood income quintile** |  |  |  |  |
| Q1 (lowest income) | 31.0 (18,779) | Ref | Ref |  |
| Q2 | 37.1 (24,559) | 1.20 (1.18, 1.22) | 1.24 (1.23, 1.26) | <.0001 |
| Q3 | 44.4 (30,350) | 1.43 (1.41, 1.45) | 1.46 (1.44, 1.49) | <.0001 |
| Q4 | 46.9 (40,697) | 1.52 (1.49, 1.54) | 1.59 (1.57, 1.61) | <.0001 |
| Q5 (highest income) | 53.3 (51,044) | 1.72 (1.70, 1.74) | 1.82 (1.80, 1.85) | <.0001 |
| **Place of residence** |  |  |  |  |
| Metro | 52.1 (103,277) | Ref | Ref |  |
| Moderate metro influence | 46.7 (27,994) | 0.90 (0.89, 0.90) | 0.81 (0.80, 0.81) | <.0001 |
| Urban | 36.3 (12,933) | 0.70 (0.69, 0.71) | 0.92 (0.89, 0.95) | <.0001 |
| Moderate urban influence | 23.7 (2,694) | 0.45 (0.44, 0.47) | 0.62 (0.59, 0.65) | <.0001 |
| Rural centre area | 32.7 (4,210) | 0.63 (0.61, 0.64) | 0.75 (0.72, 0.77) | <.0001 |
| Rural | 24.2 (12,500) | 0.46 (0.46, 0.47) | 0.63 (0.61, 0.64) | <.0001 |
| Rural Remote | 22.5 (1,821) | 0.43 (0.41, 0.45) | 0.68 (0.65, 0.72) | <.0001 |
| **Geographic health zone** |  |  |  |  |
| Calgary | 51.5 (74,339) | Ref | Ref |  |
| Edmonton | 48.9 (60,784) | 0.95 (0.94, 0.96) | 0.95 (0.94, 0.95) | <.0001 |
| South | 32.0 (8,845) | 0.62 (0.61, 0.63) | 0.88 (0.85, 0.91) | <.0001 |
| Central | 26.7 (10,247) | 0.52 (0.51, 0.53) | 0.71 (0.69, 0.73) | <.0001 |
| North | 26.0 (11,214) | 0.50 (0.50, 0.51) | 0.59 (0.58, 0.61) | <.0001 |
| **School authority type** |  |  |  |  |
| Public | 43.5 (110,057) | Ref | Ref |  |
| Publicly-funded Catholic | 47.2 (41,694) | 1.08 (1.07, 1.09) | 1.03 (1.02, 1.04) | <.0001 |
| Private | 28.8 (6,344) | 0.65 (0.65, 0.68) | 0.66 (0.64, 0.67) | <.0001 |
| Charter | 62.6 (3,602) | 1.44 (1.41, 1.47) | 1.18 (1.16, 1.21) | <.0001 |
| Francophone | 43.3(2,412) | 0.97 (0.97, 1.03) | 0.95 (0.93, 0.98) | 0.0027 |
| ECS Private Operator | 36.7 (578) | 0.84 (0.79, 0.90) | 0.87 (0.82, 0.93) | <.0001 |

