## Supplemental Files for "Coverage and correlates of COVID-19 vaccination among children aged 5-11 years in Alberta, Canada"

**Appendix**

Children born between 01 Jan 2010 and Nov 26, 2016, enrolled in AB schools in school year 2021/2022, N=382,817

Alberta children aged 5-11 years enrolled in schools

N=379,289

Exclude:

Non-Albertan children and

Children from Lloydminster, n=3,528

Alberta children aged 5-11 years included in analysis

N=377,753

Exclude:

Died or departed between September 2021 and 26 June 2022, n=1,010

Received COVID vaccine before 26 Nov 2021, n=282

Interval between two doses <21 days, n=244

**Fig A1. Cohort selection flow chart**

**Table A1. Monthly increase in vaccine coverage by age since November 26, 2021**

| Month of vaccine availability | Absolute difference in vaccine coverage between two consecutive months | | | | | | | |
| --- | --- | --- | --- | --- | --- | --- | --- | --- |
|  | Overall | 5 years (%) | 6 years (%) | 7 years (%) | 8 years (%) | 9 years (%) | 10 years (%) | 11 years (%) |
| 1st month (Nov26-Dec 26 2021) | 29.95 | 24.84 | 25.71 | 27.86 | 29.74 | 31.82 | 34.12 | 35.66 |
| Second month (Dec 27 2021-Jan 26 2022) | 9.14 | 9.12 | 8.91 | 8.68 | 8.86 | 9.09 | 8.74 | 10.75 |
| Third month (Jan 27-Feb 26 2022) | 2.98 | 2.91 | 2.86 | 2.79 | 2.97 | 2.91 | 2.88 | 3.59 |
| Fourth month (Feb 27-March 26 2022) | 0.6 | 0.56 | 0.49 | 0.56 | 0.54 | 0.63 | 0.57 | 0.89 |
| Fifth month (March 27-April 26 2022) | 0.52 | 0.47 | 0.46 | 0.42 | 0.45 | 0.42 | 0.55 | 0.85 |
| Sixth month  (April 27-May 26 2022) | 0.36 | 0.26 | 0.29 | 0.29 | 0.31 | 0.33 | 0.34 | 0.74 |
| Seventh month (May 27-June 26 2022) | 0.24 | 0.19 | 0.19 | 0.22 | 0.18 | 0.2 | 0.25 | 0.49 |

**Table A2. Sub-group analysis among children aged 11 years at the start of the study**

| Vaccine dose | Who turned 12 years during the study period | | Who remained 11 years throughout the study period | | P-value |
| --- | --- | --- | --- | --- | --- |
|  | n | % (95% CI) | n | % (95% CI) |  |
| No dose | 11,947 | 44.5 (43.9-45.1) | 11,728 | 49.9 (49.3-50.6) | <0.0001 |
| Two dose | 11,255 | 41.9 (41.3-42.5) | 8,895 | 37.9 (37.2-38.5) | <0.0001 |
| One or more doses | 14,892 | 55.5 (54.9-56.1) | 11,769 | 50.1 (49.5-50.7) | <0.0001 |

**Table A3: Time interval between first and second dose, by age**

| Age | N | Mean (Days) | Std Dev (Days) | Median (Days) | Minimum (Days) | Maximum (Days) |
| --- | --- | --- | --- | --- | --- | --- |
| 5 years | 14,170 | 67.0 | 22.4 | 61 | 21 | 210 |
| 6 years | 15,562 | 67.2 | 22.4 | 62 | 21 | 210 |
| 7 years | 16,807 | 67.0 | 22.5 | 62 | 21 | 210 |
| 8 years | 17,628 | 67.0 | 22.7 | 61 | 21 | 210 |
| 9 years | 18,905 | 67.1 | 23.0 | 61 | 21 | 210 |
| 10 years | 19,751 | 67.1 | 22.6 | 62 | 21 | 206 |
| 11 years | 20,150 | 67.4 | 24.3 | 62 | 21 | 209 |

**Table A4. COVID-19 vaccination status by selected background characteristics**

| **Characteristic** | **No doses** | | **One dose** | | **Two doses** | |
| --- | --- | --- | --- | --- | --- | --- |
|  | **n** | **% (95%CI)** | **n** | **% (95%CI)** | **n** | **% (95%CI)** |
| **Age** | | | | | | |
| 5 years | 31,427 | 61.65 (61.22-62.07) | 5,383 | 10.56 (10.29, 10.83) | 14,170 | 27.80 (27.41-28.18) |
| 6 years | 33,666 | 61.09 (60.68-61.49) | 5,884 | 10.68 (10.42, 10.93) | 15,562 | 28.24 (27.86-28.61) |
| 7 years | 33,028 | 59.18 (58.78-59.59) | 5,971 | 10.70 (10.44, 10.96) | 16,807 | 30.12 (29.74-30.50) |
| 8 years | 31,186 | 56.95 (56.54-57.37) | 5,942 | 10.85 (10.59, 11.11) | 17,628 | 32.19 (31.80-32.59) |
| 9 years | 30,299 | 54.60 (54.18-55.01) | 6,291 | 11.34 (11.07, 11.60) | 18,905 | 34.07 (33.67-34.46) |
| 10 years | 29,043 | 52.55 (52.13-52.97) | 6,474 | 11.71 (11.45, 11.98) | 19,751 | 35.74 (35.34-36.14) |
| 11 years | 23,675 | 47.03 (46.60-47.47) | 6,511 | 12.94 (12.64, 13.23) | 20,150 | 40.03 (39.60-40.46) |
| **Sex** | | | | | | |
| Female | 103,030 | 56.00 (55.77-56.23) | 20,406 | 11.09 (10.95, 11.23) | 60,547 | 32.91 (32.69-33.12) |
| Male | 109,225 | 56.42 (56.20-56.64) | 22,029 | 11.38 (11.24, 11.52) | 62,345 | 32.20 (32.00, 32.41) |
| **Neighborhood income quintile** | | | | | | |
| Lowest income (Q1) | 41,873 | 69.04 (68.67-69.41) | 6,572 | 10.84 (10.59, 11.08) | 12,207 | 20.13 (19.81, 20.45) |
| Q2 | 41,573 | 62.86 (62.50-63.23) | 7,334 | 11.09 (10.85, 11.33) | 17,225 | 26.05 (25.71, 26.38) |
| Q3 | 38,078 | 55.65 (55.27-56.02) | 8,057 | 11.77 (11.53, 12.02) | 22,293 | 32.58 (32.23, 32.93) |
| Q4 | 46,061 | 53.09 (52.76-53.42) | 9,745 | 11.23 (11.02, 11.44) | 30,952 | 35.68 (35.36, 36.00) |
| Highest income (Q5) | 44,739 | 46.71 (46.39-47.02) | 10,748 | 11.22 (11.02, 11.42) | 40,296 | 42.07 (41.76, 42.38) |
| **Place of residence** | | | | | | |
| Metro | 94,842 | 47.87 (47.65-48.09) | 24,221 | 12.23 (12.08, 12.37) | 79,056 | 39.90 (39.69, 40.12) |
| Metro Influence | 31,974 | 53.32 (52.92-53.72) | 7,029 | 11.72 (11.46, 11.98) | 20,965 | 34.96 (34.58, 35.34) |
| Urban | 22,715 | 63.72 (63.22-64.22) | 4,069 | 11.41 (11.08, 11.74) | 8,864 | 24.87 (24.42, 25.31) |
| Moderate urban | 8,686 | 76.33 (75.55-77.11) | 872 | 7.66 (7.17, 8.15) | 1,822 | 16.01 (15.34, 16.68) |
| Rural centre area | 8,677 | 67.33 (66.52-68.14) | 1,294 | 10.04 (9.52, 10.56) | 2,916 | 22.63 (21.91, 23.35) |
| Rural | 39,140 | 75.79 (75.42-76.16) | 4,280 | 8.29 (8.05, 8.53) | 8,220 | 15.92 (15.60, 16.23) |
| Rural remote | 6,290 | 77.55 (76.64-78.46) | 691 | 8.52 (7.91, 9.13) | 1,130 | 13.93 (13.18, 14.69) |
| **Geographic Health Zone** | | | | | | |
| Calgary | 70,032 | 48.51 (48.25-48.77) | 7,824 | 12.35 (12.18, 12.52) | 56,515 | 39.15 (38.89, 39.40) |
| Edmonton | 28,099 | 73.28 (72.83-73.72) | 3,468 | 9.04 (8.76, 9.33) | 6,779 | 17.68 (17.30, 18.06) |
| South | 63,413 | 51.06 (50.78-51.34) | 14,509 | 11.68 (11.50, 11.86) | 46,275 | 37.26 (36.99, 37.53) |
| Central | 18,774 | 67.97 (67.42-68.53) | 2,720 | 9.85 (9.50, 10.20) | 6,125 | 22.18 (21.69, 22.67) |
| North | 32,006 | 74.05 (73.64-74.47) | 3,935 | 9.10 (8.83, 9.38) | 7,279 | 16.84 (16.49, 17.19) |
| **School authority type** | | | | | | |
| Public | 142,731 | 56.46 (56.27, 56.66) | 28,776 | 11.38 (11.26, 11.51) | 81,281 | 32.15 (31.97, 32.24) |
| Publicly funded Catholic | 46,710 | 52.84 (52.51, 53.17) | 10,497 | 11.87 (11.66, 12.09) | 31,197 | 35.29 (234.97, 35.60) |
| Private School | 15,675 | 71.19 (70.59, 71.79) | 1,536 | 6.98 (6.64, 7.31) | 4,808 | 21.84 (21.29, 22.38) |
| Charter | 2,151 | 37.39 (36.14, 38.64) | 798 | 13.87 (12.98, 14.76) | 2,804 | 48.74 (47.45, 50.03) |
| Francophone | 3,157 | 56.69 (55.39, 57.99) | 497 | 8.92 (8.18, 9.67) | 1,915 | 34.39 (33.14, 35.63) |
| ECS Private Operator | 999 | 63.35 (60.96, 65.73) | 166 | 10.53 (9.01, 12.04) | 412 | 26.13 (23.96, 28.29) |
